# Effects of a novel infant formula on weight gain and body composition of infants: The INNOVA 2020 study

**DOI:** 10.1101/2022.10.27.22281417

**Authors:** Julio Plaza-Diaz, Francisco Javier Ruiz-Ojeda, Javier Morales, Ana Isabel Cristina de la Torre, Antonio García-García, Carlos Nuñez de Prado, Cristóbal Coronel, Cyntia Crespo, Eduardo Ortega, Esther Marín-Pérez, Fernando Ferrerira, Gema García-Ron, Ignacio Galicia, María Teresa Santos-García-Cuéllar, Marcos Moroto, Paola Ruiz, Raquel Martín, Susana Viver-Gómez, Angel Gil

## Abstract

**Background:** Breastmilk is the most appropriate food for infants and exclusive breastfeeding is highly recommended for the first six months of life to promote adequate growth and development and lower infant morbidity and mortality. Among the best-documented benefits of breastfeeding is the reduced risk of disease and infections such as pneumonia, diarrhea and acute otitis media. Nonetheless, there are situations in which the infant cannot be breastfed; therefore, it is essential to use an appropriately designed infant formula. As current infant formulas incorporate novel ingredients to partly mimic the composition of human milk, the safety and suitability of each specific infant formula should be tested by clinical evaluation in the target population. Here, we report the results of a multicenter, randomized, blinded, controlled clinical trial that aimed to evaluate a novel starting formula on weight gain and body composition of infants up to 6 and 12 months (INNOVA 2020 study), as well as safety and tolerability. The complete protocol of this study has been previously issued.

**Study design:** 210 infants (70/group) were enrolled in the study, and completed the intervention until 12 months of age. For the intervention period, infants were divided into three groups: group 1 received the formula 1 (Nutribén® Innova1 or INN), with a lower amount of protein, and enriched in α-lactalbumin protein, and with a double amount of docosahexaenoic acid (DHA)/ arachidonic acid (ARA) than the standard formula; it also contained a thermally inactivated postbiotic (*Bifidobacterium animalis* subsp. *lactis*, BPL1™ HT). Group 2 received the standard formula or formula 2 (Nutriben® Natal or STD) and the third group was exclusively breastfed for exploratory analysis and used as a reference (BFD group). During the study, visits were made at 21 days, 2, 4, 6, and 12 months of age, with ± 3 days for the visit at 21 days of age, ± 1 week for the visit at 2 months, and ± 2 weeks for the others. During the first 6 months of the study, the infants were only supplied with the starting formula or natural breastfeeding.

**Results:** The primary outcome, weight gain, was higher in both formula groups than in the BFD group at 6 and 12 months, whereas no differences were found between STD and INN groups neither at 6 nor at 12 months. Likewise, BMI was higher in infants fed the two formulas compared with the BFD group. Regarding body composition, length, head circumference and tricipital/subscapular skinfolds were alike between groups. The INN formula was considered safe as weight gain and body composition were within the normal limits, according to WHO standards. The BFD group exhibited more liquid consistency in the stools compared to both formula groups. All groups showed similar digestive tolerance and infant behavior. However, a higher frequency of gastrointestinal (GI) symptoms was reported by the STD formula group (291), followed by the INN formula (282) and the BFD groups (227). There were fewer respiratory, thoracic, and mediastinal disorders among BFD children. Additionally, infants receiving the INN formula experienced significantly fewer general disorders and disturbances than those receiving the STD formula. Indeed, atopic dermatitis, bronchitis, and bronchiolitis were significantly more prevalent among infants who were fed the STD formula compared to those fed INN formula or breastfed. To evaluate whether there are significant differences between formula treatments, beyond growth parameters, it would seem necessary to examine more precise health biomarkers and to carry out long-term longitudinal studies.

**Clinical Trial Registration:** The trial was registered with Clinicaltrial.gov (NCT05303077) on March 31, 2022, and lastly updated on April 7, 2022.

## 1. Introduction

Exclusive breastfeeding is the gold standard for infant feeding because it promotes adequate growth and development, excellent nutritional status, and appropriate psychological development. In addition, due to the special composition of breast milk in bioactive and immunogenic substances, it effectively protects against numerous infectious diseases, mainly pneumonia and other respiratory infections, diarrhea, and allergic processes. Moreover, breastfeeding promotes an optimal psycho-affective mother-child relationship and has a very low cost, practically nil (Ip et al., 2009;Walker, 2010;Black et al., 2013;Victora et al., 2016;Ogbu et al., 2021). Indeed, WHO and UNICEF recommend starting breastfeeding within the first hour of birth and being exclusively breastfed for the first 6 months of life (World Health Organization, 2002;2022). However, from the age of 6 months, children should begin eating safe and adequate complementary foods while continuing to breastfeed for up to 2 years and beyond (Kramer and Kakuma, 2001).

Breastfeeding protects against disease in both developing (Black et al., 2013) and developed countries (Ip et al., 2009) and is beneficial not only for infants but also for mothers. Indeed, breastfeeding may protect later in life against obesity and metabolic diseases (Rameez et al., 2019;World Health Organization, 2022). Additionally, breastfeeding is associated with better performance on intelligence tests (Horta et al., 2015;Victora et al., 2015). Furthermore, women who breastfeed have a reduced risk of breast and ovarian cancers (World Health Organization, 2022).

Despite worldwide efforts to promote breastfeeding, there are many social factors, especially premature return to work after childbirth and, in some cases, maternal illness, which cause many mothers to abandon breastfeeding prematurely (Balogun et al., 2015). To promote the adequate growth and development of general infants and those who have those conditions, infant formula designed with an optimal nutritional composition is essential. In this regard, different entities such as the European Society for Paediatric Gastroenterology, Hepatology and Nutrition (ESPGHAN) (Koletzko et al., 2005) and the American Academy of Paediatrics (AAP) have established various recommendations for the composition of infant formulas. In addition, various governmental organizations, such as the FAO/WHO Codex Alimentarius Commission (CODEX ALIMENTARIS, 2020), the European Union Commission (Commission Delegated Regulation (EU) 2016) and the US Food and Drug Administration (FDA) (FOOD AND DRUG ADMINISTRATION, 2018) have established strict compositional and control standards for infant formula foods to protect the health of consumers and ensure fair practices in the food trade.

Continuous research on the composition of human milk and the biological effects of its components (Christian et al., 2021) has led to the constant evolution of infant formula, especially during the last five decades, incorporating various food ingredients to meet not only the nutritional needs of infants but also to contribute to better development and functionality (Carver, 2003). Thus, based on certain studies suggesting that a high protein intake in the early stages of life may be the cause of obesity and increased risk of metabolic disease in later stages of life (Brands et al., 2014;Zheng et al., 2018), the protein composition of infant formulas has been adjusted both in quality and quantity, reducing the protein intake of infants (Weber et al., 2014; Totzauer et al., 2018;Kouwenhoven et al., 2021). Regarding the quality of protein intake, the whey/casein ratio in infant formulas is important for the first year of life. Whey proteins, both β-lactoglobulin and α-lactalbumin, are rapidly digested and participate in the building of muscle mass (Boirie et al., 1997), being the α -lactalbumin the major protein in human milk (Liao et al., 2017). Therefore, development of infant formula containing bovine α-lactalbumin may improve the plasma amino acid pattern of the receiver infant, allowing a reduction in the protein content of the formula. On the other hand, the relatively high content of long-chain polyunsaturated fatty acids of both the n-6 and n-3 series, especially arachidonic acid (AA, 20:4 n-6) and docosahexaenoic acid (DHA, 22:6 n-3) in human milk and their proven effects on the cognitive development of infants, has led to the incorporation of these fatty acids into infant formulas. In this regard, and even though EFSA and the European Union Commission have established mandatory compositional recommendations only for DHA content (EFSA Panel on Dietetic Products and Allergies, 2014;EU Commission, 2016), numerous researchers have argued and established consensus documents on the need for infant formulas to incorporate both AA and DHA in amounts comparable to the average composition of human milk. Hence, according to some studies infant formula should provide DHA at levels of 0.3% to 0.5% by weight of total fatty acids and with a minimum level of ARA equivalent to the DHA content (Koletzko et al., 2020;Campoy et al., 2021). Therefore, the supplementation of infant formulas with both DHA and ARA in appropriate amounts should be clinically tested.

During the last two decades, advances in the knowledge of the composition of the intestinal microbiome of breast-fed infants and its beneficial effects on the host (Flaherman et al., 2018;Gopalakrishna and Hand, 2020) have led to the incorporation of both prebiotics and probiotics in infant formulas (Thomas et al., 2010;Braegger et al., 2011). In this regard, improvements in the prevention of acute infectious diarrhea, treatment of antibiotic-associated diarrhea and prevention of allergy have been reported (Thomas et al., 2010;Braegger et al., 2011;Szajewska et al., 2016;Adjibade et al., 2022). Also, it is known that not only live probiotics but also their fermentation products and death cells derived from inactivated probiotics, referred to by the International Scientific Association of Probiotics and Prebiotics (ISAPP) as postbiotics, can exert biological effects of interest on the intestinal microbiota (Salminen et al., 2021;Vinderola et al., 2022). In this context, *Bifidobacterium animalis* subsp. *lactis* CECT 8145 inactivated by heat (BPL1™ HT) exhibits several biological effects of interest such as reduction in fat deposition via the IGF-1 pathway in mice (Balaguer et al., 2022) and has anti-obesity effects in obese Zucker rats (Carreras et al., 2018). Moreover, daily consumption of the living strain BPL1™ ameliorates several anthropometric adiposity biomarkers in abdominally obese adults (Pedret et al., 2019). Likewise, an infant formula supplemented with BPL1™ HT reduced fat deposition in *C. elegans* and augmented acetate and lactate in a fermented infant slurry (Silva et al., 2020).

Novel infant formulas should be thoroughly evaluated to ensure their safety, efficacy, and tolerability. Hence, in the present study, we aimed to evaluate the effect of a novel starting infant formula on weight gain, body composition, safety, and tolerability, in infants up to 6 and 12 months of age. This was compared with standard infant formula. Safety and tolerability outcomes included digestive tolerance (flatulence, vomiting and regurgitation), stool appearance (consistency and frequency), behavior (restlessness, colic, nocturnal awakenings), and incidence of infections. Safety objectives included any frequency of GI symptoms resulting from the consumption of the study formula. As a further exploratory objective, we compared weight gain up to 6 and 12 months and changes in the other secondary outcomes among the infants who received the modified starter formula and the standard formula with a group of exclusively breastfed infants. The complete design and methodology of this randomized, multicenter, double-blind, parallel, comparative clinical trial has been previously registered with Clinicaltrial.gov (NCT05303077) on March 31, 2022, and lastly updated on April 7, 2022, and published (Ruiz-Ojeda et al., 2022).

## 2. Material and methods

### 2.1. Ethics

This clinical trial was carried out following the recommendations of the International Conference on Harmonization Tripartite on PCBs, the ethical-legal principles established in the latest revision of the Declaration of Helsinki, as well as the current regional regulations that regulate pharmacovigilance and food safety. The present study was approved by the Committee for Technical Investigation in Regional Medicine in the Madrid Community (CEIm-R) dated 11/05/2018 under the name INNOVA2020 version 2.0. All personal data obtained in this study are confidential. They were treated under the Spanish Organic Law 3/2018, of December 5, on the Protection of Personal Data and guarantee of digital rights. The researchers or the institutions implicated in the study allowed direct access to the data or source documents for monitoring, auditing, and review by the CEIm-R. They also allowed inspection of the trial by the health authorities.

### 2.2. Trial design

The INNOVA study was designed as a randomized, multi-center, double-blind, parallel, and comparative clinical trial of the equivalence of two starting infant formulas for infants. Furthermore, a third un-blinded group of breastfed infants was used as a further reference group for exploratory analysis. Blinding for both investigator and participant remained assured as both infant formulas were labeled the same. It is not mandatory to carry out specific clinical tests to demonstrate the nutritional and healthy properties of infant formulas as per the current EU legislation (EC Regulation No. 1924/2006) (Ruiz-Ojeda et al., 2022).

Parents were informed by pediatricians involved in the study of the possibility of participating in it at a meeting at 15 days of infant age. After accepting to participate, they came to the health center for the baseline visit a week later (visit at 21 days of age). The pediatrician requested, at the meeting at 15 days of life, that parents provide at the next visit relevant information from the maternal history that was required for the study and that was not available in the pediatric history. The visit that took place within the scheduled time was considered valid with a margin of ± 3 days in the visit at 21 days of age, ± 1 week in the visit at 2 months of age, and ± 2 weeks in visits at 4, 6 and 12 months of age. The inclusion criteria were healthy children of both sexes; term children (between 37 and 42 weeks of gestation), birth weight between 2500-4500 g, single delivery, and mothers with a body mass index, before pregnancy, between 19 and 30 kg/m^2^. The exclusion criteria were, body weight less than the 5^th^ percentile for that gestational age at the time of the inclusion, allergy to cow’s milk proteins and/or lactose, history of antibiotic use during the 7 days before inclusion, congenital disease or malformation that can affect growth, diagnosis of disease or metabolic disorders, significant prenatal and/or severe postnatal disease before enrollment, minor parents (younger than 18 years old), newborn of a diabetic mother, newborn of a mother with drug dependence during pregnancy, newborn whose parents/caregivers cannot comply with procedures of the study, and infants participating or have participated in another clinical trial since their birth. Participants had the right to withdraw from the study at any time and for any reason, without giving explanations or suffering any penalty for it. Likewise, the investigator could withdraw study participants if they did not comply with the study procedures. This is for any reason that, in the investigator’s opinion, may pose the infants at risk or makes it necessary to suspend breastfeeding. Children who received antibiotics during the 7 days before inclusion were not considered eligible for the study and, children who received antibiotics during the trial, were excluded: Any treatment, food or product that could interfere with the trial at any time, and breasfeeding which was different from that of the group assigned group in the trial.

The sample size of the trial is 210 children (70/group), based on the main outcome of weight gain, which is the main variable chosen according to the guidelines “Guidelines from the American Academy of Pediatrics Task Force on Clinical Testing of Infant Formulas” (Task force on clinical testing of infant formulas committee on nutrition. American Academy of Pediatrics Committee on Nutrition et al., 1988). The infants were selected by Primary Care pediatricians through active and consecutive recruitment i.e. pediatricians informed and invited parents of 15-day-old infants not being breasfed for some reasons to be involved in the trial (see details below under item 2.3). The study was carried out in 21 centers, all located in Spain, of which 17 recruited at least one subject. In total, 217 subjects signed the informed consent (IC) and 145 were randomized to receive one of the two infant formulas. Of these 145, 3 were randomization failures and 2 were screening failures, thus 140 infants who met all the inclusion criteria and no exclusion criteria were included in the study, and 70 were unblinded within infants exclusively breastfed. A total of 185 subjects completed all study visits, of which there were 25 dropouts, 12 in the breastfeeding group, 8 in the formula 1 group, and 5 in the standard formula group. The trial was registered with clinicaltrial.gov website (NCT05303077, https://clinicaltrials.gov/ct2/show/NCT05303077) on March 31, 2022, and lastly updated on April 7, 2022 (Ruiz-Ojeda et al., 2022). The inclusion period started in October 01, 2018; and the follow-up period was between February 11, 2019 and November 25, 2020.

### 2.3 Formula characteristics

- Group 1 (Infant formula 1): Nutribén^®^ Innova 1
- Group 2 (Infant formula 2): Nutribén® Standard
- Group 3: Breastfeeding (External control exploratory analysis)

Infants were recruited from Primary Care Pediatrics clinics by the pediatricians participating in the trial. Pediatricians informed and invited parents of 15-day-old infants who visited their offices regularly (for regular medical check-ups) to be involved in the trial. Infants that not receive breastfeeding at the time of inclusion (for different reasons) were proposed to participate in the formula-feeding groups. To keep the three arms of the trial balanced, one candidate breastfeeding subject was recruited at each center for every two infants supplemented with infant formula. The selection of the children was made among those infants who met the inclusion and exclusion criteria of the study and fit the categories of sex, body mass index (BMI) of the mother before pregnancy (<25 kg/m^2^ or >25 kg/m^2^), and birth weight (<3500 g or >3500 g).

The experimental product object of this trial (Formula 1 or Nutribén® Innova 1, INN formula) and the Nutribén® Standard formula (STD formula) comply with the recommendations of the ESPGHAN (European Society of Pediatric Gastroenterology, Hepatology and Nutrition) and with Regulation 609/2013 of the European Parliament and of the Council regarding foods intended for children, infants and young children, foods for special medical purposes and complete diet substitutes for weight control and repealing Council Directives 92/51, Directives 96/8/EC, 1999/21/CE, 2006/125/CE and 2006/141/CE of the Commission, Directive 2009/38/CE of the European Parliament and of the Council and Regulations 41/2009 and 953/2009 of the Commission. More detailed information on the composition of each of the products can be found in Table 1. Infant formulas are given ad libitum orally. The two trial formulations were administered following the preparation instructions in the manufacturer’s package insert. DHA was obtained from purified and concentrated fish oil.

**Table 1.** Nutritional composition of the standard infant formula (STD) and study formula (INN).

| Composition | STD formula |  |  | INN formula |  |  |
| --- | --- | --- | --- | --- | --- | --- |
|  | 100 g | 100 mL | 100 kcal | 100 g | 100 mL | 100 kcal |
| Energy (kcal) | 514 | 67 |  | 489 | 67 |  |
| Energy (kJ) | 2152 | 280 |  | 2046 | 279 |  |
| Total Fat (g) | 27 | 3.5 | 5.2 | 25.6 | 3.5 | 5.2 |
| Linoleic acid (□-6) (mg) | 3.79 | 493 | 736 | 3.147 | 427 | 641 |
| α-linolenic acid (□-3) (mg) | 500 | 65 | 97 | 387 | 53 | 79 |
| Arachidonic acid AA (□-6) (mg) | 53 | 6.9 | 10 | 118 | 16 | 24 |
| Docosahexaenoic acid DHA (□-3) (mg) | 53 | 6.9 | 10 | 118 | 16 | 24 |
| Carbohydrates (g) | 56.4 | 7.3 | 11 | 54.6 | 7.4 | 11.1 |
| Total sugars (g) | 56.4 | 7.3 | 11 | 53 | 7.2 | 10.8 |
| Lactose (g) | 55 | 7.2 | 10.7 | 53 | 7.2 | 10.8 |
| Galacto-oligosaccharides (g) | 3.1 | 0.4 | 0.6 | 2.9 | 0.4 | 0.6 |
| Proteins (g) | 10.6 | 1.4 | 2.1 | 9.4 | 1.3 | 1.9 |
| Whey proteins (g) | 6.4 | 0.83 | 1.2 | 6.6 | 0.9 | 1.3 |
| Caseins (g) | 4.2 | 0.55 | 0.82 | 2.8 | 0.38 | 0.57 |

### 2.4 Measurements and evaluations

#### 2.4.1 Anthropometric measures

Weight, length, head circumference, body mass index, body fat percentage, and lean body mass measurements were taken at all study visits (*visit 1*, 21 days, *visit 2*, 2 months, *visit 3*, 4 months, *visit 4*, 6 months, and *visit 5*, 12 months). Fat mass was estimated by the skinfold measurements. From the visit scheduled for the fourth month of life, the mean arm circumference, the triceps skinfold, and the subscapular skinfold were added to the previous measurements. All these measurements were done in duplicate. Using as valid data the mean between the two. Each center had all the necessary materials to carry out these measurements. They needed to always use the same material with all subjects and, of course, the material should be calibrated.

#### 2.4.2 Characteristics of feces and digestive tolerance

We evaluated consistency and the number of stools/day) at times 21 days, 2, 4, 6 and 12 months of age Flatulence, vomiting and regurgitation at times 21 days, 2, 4, 6 and 12 months of age were also assessed.

#### 2.4.3 Infant’s behavior evaluated by parents

Restlessness, colic, and night awakenings were evaluated during 21 days, 2, 4, 6 and 12 months of age for the parents. More details are available in the study protocol (Ruiz-Ojeda et al., 2022).

#### 2.4.4 Morbidity, safety and tolerability

Variables related to morbidities were evaluated within 1-year. All morbidities were coded according to MedDRA dictionary version 21.0 (MedDRA, 2018).

### 2.5 Statistical methods

The main objective of the study was to evaluate whether the weight gain was equivalent between treatment groups receiving formula 1 and 2, and it has been decided that the sample size of the trial is 210 children (70/group) based on the primary variable weight gain (the primary variable selected), following the “Guidelines from the American Academy of Pediatrics” issued by the Task Force on Clinical Testing of Infant Formulas of the American Academy of Pediatrics (Task force on clinical testing of infant formulas committee on nutrition. American Academy of Pediatrics Committee on Nutrition et al., 1988). Previous studies carried out on children fed from 0 to 6 months with different formulations of infant formula have shown a mean weight gain of around 20-25 g/day with a standard deviation between 5 and 6 g/day (Bosheva et al., 2022). In most of these studies, a difference in mean weight gain of 3 g/day was considered clinically relevant. Thus, the main objective was resolved by using a t-test for independent samples. Considering a power of 80%, a significance level of 5%, an equivalence limit of 3 g/day, and a common standard deviation of 5.5 g/day, we needed to recruit at least 59 children in each of the groups. Based on an estimated dropout rate of 20%, it was necessary to include 70 children in each of the groups, which means 140 infants. A third group of the same size (70 children) was included for the secondary comparisons between the bottle-fed and breastfed groups, maintaining the same significance and power in this secondary comparison as in the main comparison. Categorical variables are described as frequencies or percentages. Continuous variables are reported as mean and standard deviation. Between-group differences in measures of growth and body composition are analyzed using analysis of variance, including analysis of covariance and general models of analysis of variance for repeated measures (GLM-ANOVA and MANOVA) when required. The Chi-square test is applied to compare discrete variables between groups. The Bonferroni correction is used when comparisons are made between more than two groups. The alpha level of significance is set at 0.05. All evaluable infants have been included in the statistical analysis, considering “evaluable” all those who have the main measurement variable. Both, intention to treat and protocol-based statistical analyses are performed.

## 3. Results

**Table 2** shows demographic and descriptive information for infants and their mothers. Here, gestational age, birth weight, childbirth delivery, assisted reproductive technology, and the mother’s medical background were included, though no significant variables were shown. During visit 1 (21 days), only daily depositions per day were significantly higher for the BFD group in comparison with formula groups.

**Table 2.** Baseline demographics of the infants in the study

|  | **BFD** | **STD** | **INN** | **P-value** |
| --- | --- | --- | --- | --- |
| **N** | 70 | 70 | 70 |  |
| **Gender (% of women)** | 28 (40.0) | 29 (41.4) | 33 (47.1) | 0.738 |
| **Gestational age (weeks)** | 39.44 (1.1) | 39.20 (1.1) | 39.34 (0.9) | 0.389 |
| **Birth weight (g)** | 3359.20 (403.9) | 3300.87 (462.9) | 3363.30 (408.7) | 0.626 |
| *Birth weight >3500 g (%)* | 22 (31.4) | 22 (31.4) | 27 (38.6) | 0.613 |
| **Childbirth delivery n, (%)** |  |  |  | 0.579 |
| *Cesarean section* | 13 (18.6) | 17 (24.3) | 20 (28.6) |  |
| *Distocyc* | 8 (11.4) | 6 (8.6) | 9 (12.9) |  |
| *Normal childbirth* | 49 (70.0) | 47 (67.1) | 41 (58.6) |  |
| **Assisted reproductive technology n, (%)** | 4 (5.7) | 6 (8.6) | 3 (4.3) | 0.679 |
| *Assisted reproductive technology type (%)* |  |  |  | 0.066 |
| *In vitro fertilization* | 1 (25.0) | 6 (100.0) | 2 ( 66.7) |  |
| *Artificial insemination* | 2 (50.0) | 0 (0.0) | 1 ( 33.3) |  |
| *Egg donation* | 1 (25.0) | 0 (0.0) | 0 (0.0) |  |
| **Maternal BMI (kg/m<sup>2</sup>), (x, SD)** | 23.72 (3.2) | 23.92 (3.3) | 23.78 (3.0) | 0.967 |
| *BMI mother >25 n, (%)* | 22 ( 31.4) | 22 ( 31.4) | 24 ( 34.3) | 0.85 |
| **Mother's medical background n, (%)** |  |  |  | 0.849 |
| *Allergy* | 0 (0.0) | 0 (0.0) | 2 (2.9) |  |
| *Hypothyroidism* | 7 (10.0) | 5 (7.1) | 4 (5.7) |  |
| *Arterial hypertension* | 0 (0.0) | 2 (2.9) | 1 (1.4) |  |
| *No* | 58 (82.9) | 55 (78.6) | 55 (78.6) |  |
| *Others* | 5 (7.1) | 8 (11.4) | 8 (11.4) |  |
| **Visit 1, (21 days)** | **BFD** | **STD** | **INN** | **P-value** |
| **Depositions per day** | 5.2 (3.2) | 2.5 (2.0) | 2.3 (1.4) | <0.001* |
| **Weight** | 3921.8 (475.5) | 3805.0 (507.5) | 3916.6 (453.9) | 0.268 |
| **Length** | 53.1 (1.9) | 52.6 (2.2) | 52.8 (1.7) | 0.287 |
| **Head circumference** | 36.6 (1.0) | 36.2 (1.3) | 36.5 (1.2) | 0.214 |

Data are expressed as mean and standard deviation, and frequency or percentage. BMI, body mass index; NS, non-significant

The parents or legal guardians returned to the center all the leftover containers (empty containers and containers with products) to record the amount of product dispensed and the amount of infant formula used up to the time of the visit; the corresponding weighing was performed and the percentage of product ingested by the participants was determined. Finally, the grams of formulation consumed by each infant, measured by weighing the leftover formulations or as indicated in the parents’ diary corresponding to the periods between visits 1-2, 2-3 and 3-4, were available. The participants of this study included in the formula-feeding groups were considered compliant if they took at least 80% of the formula during the first six months while in the breastfeeding group it was considered a valid external control if more than 80% of the feedings were breastfeeding. All infants included in the BFD group, except one, consumed more than 80% of human milk during the first six months.

Based on the record of the amount of product dispensed and the amount of infant formula used (measurement of leftover) we estimated the average daily intake from visits 1 to 4 for the STD group (130.2 ± 14.7 g/d, equivalent to 671±76 kcal/d of energy and 13.8 ± 1.56 g/d of protein) and the INN group (134.0 ±18.2 g/d, equivalent to 655 ± 89 kcal /d of energy and 12.60 g/d of protein).

A GLM-ANOVA was performed for grams of formula consumed per day, and significant differences were observed for visit, sex and infant birth weight, but not about the formula used (data not presented).

The primary variable (weight gain) consisted of the difference in infant weight (in g/day) between the initial recruitment visit and the 6-month visit, divided by the number of days between the two visits. This analysis was performed on the 187 infants who completed until visit 4 (6 months). The difference between visit 1 and visit 4 was calculated in days of 187 children individually (**Table 3**). When analyzing the mean difference in weight gain between the groups of children treated with the STD and INN formulations (**Table 3**), there was no statistically significant difference in weight gain between the two formulations. On the other hand, an ANOVA of the weight difference between visits 1 and 4 was performed for the STD and INN formulations controlling for the factors, gender, maternal body mass index (BMI) and birth weight. There were significant differences in weight difference between visits 1 and 4 as a function of gender and infant birth weight, but not as a function of formulation or mother’s BMI (**Table 3**). The grams consumed per day were also related to the difference in weight between visits 1 and 4. Based on the weighing of leftover grams of the dispensed formulation, a multivariate analysis of variance (MANOVA) of the difference in weight of the children between visits 1 and 4 was performed as a function of formulation consumed (STD, INN), gender (male, female), maternal BMI (<25, >25), birth weight (<3500, >3500) and grams ingested per day. A moderate positive correlation (r=0.535) was observed between mean grams ingested per day (measured by weighing leftover formulations) and weight gain between visits 1 and 4 of the infants. This correlation changed significantly by gender (males achieved larger weight differences for the same daily grams of formulation consumed) and by birth weight (infants with birth weights > 3500 g achieved larger weight differences for the same daily grams of formulation consumed) but not as a function of maternal BMI or formulation used.

**Table 3.**
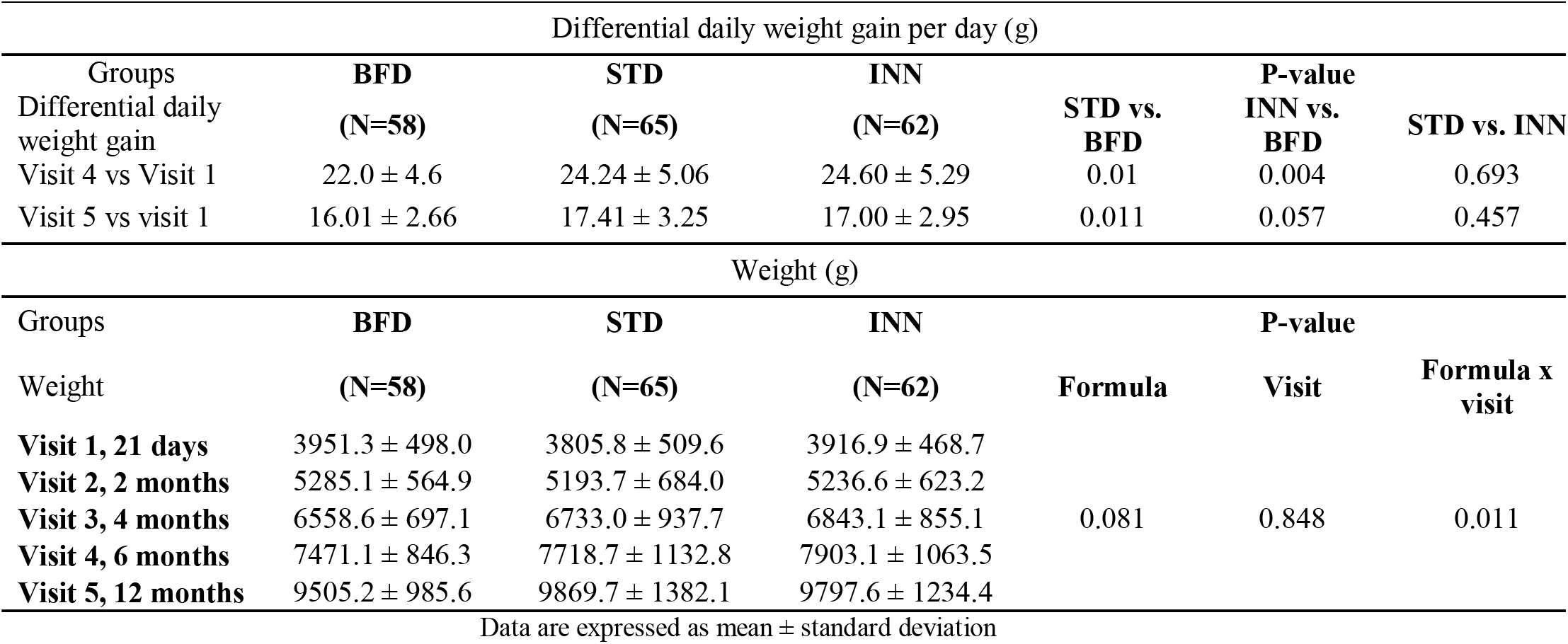
Differential daily weight gain and absolute weight in the entire study

| Differential daily weight gain per day (g) |  |  |  |  |  |  |
| --- | --- | --- | --- | --- | --- | --- |
| Groups | BFD | STD | INN |  | P-value |  |
| Differential daily weight gain | (N=58) | (N=65) | (N=62) | STD vs. BFD | INN vs. BFD | STD vs. INN |
| Visit 4 vs Visit 1 | 22.0 ± 4.6 | 24.24 ± 5.06 | 24.60 ± 5.29 | 0.01 | 0.004 | 0.693 |
| Visit 5 vs visit 1 | 16.01 ± 2.66 | 17.41 ± 3.25 | 17.00 ± 2.95 | 0.011 | 0.057 | 0.457 |
| Weight (g) |  |  |  |  |  |  |
| Groups | BFD | STD | INN |  | P-value |  |
| Weight | (N=58) | (N=65) | (N=62) | Formula | Visit | Formula x visit |
| **Visit 1, 21 days** | 3951.3 ± 498.0 | 3805.8 ± 509.6 | 3916.9 ± 468.7 |  |  |  |
| **Visit 2, 2 months** | 5285.1 ± 564.9 | 5193.7 ± 684.0 | 5236.6 ± 623.2 |  |  |  |
| **Visit 3, 4 months** | 6558.6 ± 697.1 | 6733.0 ± 937.7 | 6843.1 ± 855.1 | 0.081 | 0.848 | 0.011 |
| **Visit 4, 6 months** | 7471.1 ± 846.3 | 7718.7 ± 1132.8 | 7903.1 ± 1063.5 |  |  |  |
| **Visit 5, 12 months** | 9505.2 ± 985.6 | 9869.7 ± 1382.1 | 9797.6 ± 1234.4 |  |  |  |

Data are expressed as mean ± standard deviation

This same MANOVA as a function of grams ingested daily according to the parental diary also showed a positive correlation (r=0.626) between mean grams ingested per day (obtained from parental diary entries) and weight gain between visits 1 and 4 of the infants. This correlation changed significantly by gender (boys achieved larger weight differences for the same daily grams of formulation consumed) and by birth weight (children with birth weights > 3500 achieved larger weight differences, especially those who consumed more daily grams of formulation) but not as a function of maternal BMI or formulation used.

Secondarily and foreseen in the study protocol, the weight gain of the infants who received each of the formulations studied was compared with the BFD group. The results of this analysis are shown in **Table 3**, where it can be seen that BFD infants had a significantly lower weight gain at 6 months of age than that shown by infants fed with either of the two formulations evaluated (STD and INN).

The same analysis was performed for the variable weight gain at 12 months of study. **Table 3** shows the number of children in each of the 3 groups studied with weight values recorded at visit 1 (21 days of age) and visit 5 (12 months ± 2 weeks). A comparison of the means of weight gain from the different formulations (STD vs. INN, STD vs. BFD and, INN vs. BFD, **Table 3**) revealed that at 6 months there is no significant difference in weight gain (measured in g/day) as well as at 12 months between the STD and INN formulations. The total weight of the STD formulation group remained significantly higher than the BFD group at 12 months, however, the INN group exhibited a trend to be different from breastfeeding between visits 1 and 5 (p=0.057). A GLM-ANOVA (data not presented) showed significant differences for visit (p<0.001) and for the interaction between formulation and visit (p=0.004), but not for formulation exclusively (p=0.381). The largest effect size was associated with the visit factor.

No significant differences were observed in length and head circumference, as well as, tricipital and subscapular skinfolds and mean upper arm circumference (**Tables 4** and **5**).

**Table 4.** Length and head circumference in the entire study

| Visits | Length (cm) |  |  |
| --- | --- | --- | --- |
|  | BFD<br>(N=58) | STD<br>(N=65) | INN<br>(N=62) |
| **Visit 1** | 53.15 ± 2.08 | 52.49 ± 2.24 | 52.87 ± 1.78 |
| **Visit 2** | 57.89 ± 2.42 | 57.47 ± 2.33 | 57.95 ± 2.02 |
| **Visit 3** | 63.16 ± 2.42 | 63.24 ± 3.67 | 63.50 ± 2.16 |
| **Visit 4** | 66.68 ± 2.64 | 67.01 ± 3.26 | 67.15 ± 2.14 |
| **Visit 5** | 75.44 ± 2.90 | 75.82 ± 3.08 | 75.96 ± 2.54 |
| Visits | Head circumference (cm) |  |  |
|  | BFD<br>(N=58) | STD<br>(N=65) | INN<br>(N=62) |
| **Visit 1** | 36.66 ± 1.02 | 36.22 ± 1.34 | 36.51 ± 1.25 |
| **Visit 2** | 36.09 ± 1.17 | 38.87 ± 1.36 | 39.11 ± 1.63 |
| **Visit 3** | 41.56 ± 1.12 | 41.79 ± 3.73 | 41.64 ± 1.39 |
| **Visit 4** | 43.19 ± 1.22 | 43.18 ± 1.37 | 43.41 ± 1.45 |
| **Visit 5** | 46.40 ± 1.18 | 46.25 ± 1.53 | 46.29 ± 1.50 |

Data are expressed as mean and standard deviation

**Table 5.** Tricipital and subscapular skinfolds and mean upper arm circumference in visits 3, 4, and 5.

| Tricipital skinfold (mm) |  |  |  |
| --- | --- | --- | --- |
|  | **BFD** | **STD** | **INN** |
|  | **(N=58)** | **(N=65)** | **(N=62)** |
| **Visit 3** | 12.32 ± 2.26 | 12.66 ± 2.67 | 12.78 ± 3.00 |
| **Visit 4** | 12.35 ± 2.23 | 12.60 ± 2.65 | 12.90 ± 2.99 |
| **Visit 5** | 12.34 ± 2.23 | 12.63 ± 2.65 | 12.84 ± 2.99 |
| Subscapular skinfold (mm) |  |  |  |
|  | **BFD** | **STD** | **INN** |
|  | **(N=58)** | **(N=65)** | **(N=62)** |
| **Visit 3** | 9.92 ± 2.69 | 9.83 ± 2.72 | 10.25 ± 2.73 |
| **Visit 4** | 9.90 ± 2.49 | 9.94 ± 2.72 | 10.36 ± 2.73 |
| **Visit 5** | 9.91 ± 2.58 | 9.89 ± 2.73 | 10.30 ± 2.72 |
| Mean upper arm circumference (cm) |  |  |  |
|  | **BFD** | **STD** | **INN** |
|  | **(N=58)** | **(N=65)** | **(N=62)** |
| **Visit 3** | 13.79 ± 1.30 | 13.89 ± 1.67 | 14.15 ± 1.21 |
| **Visit 4** | 14.41 ± 1.24 | 14.72 ± 1.46 | 14.98 ± 1.88 |
| **Visit 5** | 15.34 ± 1.26 | 15.29 ± 1.68 | 15.94 ± 1.39 |

Data are expressed as mean and standard deviation

In the case of BMI, the ANOVA (**Table 6**) shows significant differences for visit (p<0.001) and for the interaction between formulation and visit (p<0.001), but not for formulation (p=0.487). The largest effect size was associated with the visit factor (0.4230, data not provided). In addition, we have reported that it was from the third visit onwards that the mean BMI was higher in the STD and INN formulations compared to the BFD group; hence, the interaction effect appears as significant (**Table 6**). For the body fat percentage, the ANOVA (**Table 6**) shows significant differences for visit (p=0.005), but not for the interaction between formulation and visit (p=0.958) and for formulation (p=0.249). The largest effect size is associated with the visit factor (0.0236, data not shown). Finally for the lean mass, the ANOVA (**Table 6**) shows significant differences for visit (p<0.001), but not for the interaction between formulation and visit (p=0.054) and for formulation (p=0.215). The largest effect size is associated with the visit factor (0.6093, data not provided).

**Table 6.** Body mass index, body fat percentage, and lean mass.

| Body mass index (kg/m<sup>2</sup>) |  |  |  | **P-value** |  |  |
| --- | --- | --- | --- | --- | --- | --- |
| **Visits** | **BFD** | **STD** | **INN** | **Formula** | **Visit** | **Formula x visit** |
|  | **(N=58)** | **(N=65)** | **(N=62)** |  |  |  |
| **Visit 1** | 13.94 ± 1.11 | 13.75 ± 1.04 | 13.97 ± 1.07 |  |  |  |
| **Visit 2** | 15.74 ± 1.08 | 15.67 ± 1.30 | 15.56 ± 1.28 |  |  |  |
| **Visit 3** | 16.42 ± 1.22 | 16.97 ± 3.61 | 16.94 ± 1.59 | 0.487 | <0.001 | <0.001 |
| **Visit 4** | 16.78 ± 1.30 | 17.15 ± 1.83 | 17.49 ± 1.81 |  |  |  |
| **Visit 5** | 16.69 ± 1.28 | 17.11 ± 1.64 | 16.94 ± 1.54 |  |  |  |
| Body fat percentage |  |  |  | **P-value** |  |  |
| **Visits** | **BFD** | **STD** | **INN** | **Formula** | **Visit** | **Formula x visit** |
|  | **(N=58)** | **(N=65)** | **(N=62)** |  |  |  |
| **Visit 3** | 19.35 ± 3.53 | 19.60 ± 3.39 | 20.37 ± 3.67 |  |  |  |
| **Visit 4** | 20.36 ± 3.53 | 20.30 ± 3.73 | 21.29 ± 3.93 | 0.249 | 0.005 | 0.958 |
| **Visit 5** | 20.84 ± 3.64 | 21.02 ± 3.82 | 21.61 ± 4.01 |  |  |  |
| Lean mass (g) |  |  |  | **P-value** |  |  |
| **Visits** | **BFD** | **STD** | **INN** | **Formula** | **Visit** | **Formula x visit** |
|  | **(N=58)** | **(N=65)** | **(N=62)** |  |  |  |
| **Visit 3** | 5284 ± 559 | 5403 ± 709 | 5436 ± 615 |  |  |  |
| **Visit 4** | 5942 ± 654 | 6135 ± 823 | 6207 ± 782 | 0.215 | <0.001 | 0.054 |
| **Visit 5** | 7509 ± 703 | 7805 ± 1023 | 7670 ± 967 |  |  |  |

Data are expressed as mean and standard deviation

In the case of BMI percentiles, the ANOVA (**Table S1**) reveals significant differences for visit (p=0.009) and the interaction between formulation and visit (p=0.006), but not for formulation (p=0.504). The largest effect size is associated with the visit factor (0. 0222, data not provided). In the ANOVA, there were significant differences in the height percentiles for visit (p=0.011), for the interaction between formulation and visit (p<0.001), but not for formulation (p=0.433, **Table S1**). The largest effect size is associated with the visit factor (0.0154, data not shown). Finally, in the weight percentiles, the ANOVA (**Table S1**) shows significant differences for visit (p<0.001) and for the interaction between formulation and visit (p<0.001), but not for formulation (p=0.368). The largest effect size is associated with the visit factor (0.0228, data not provided).

The time course of the study shows a difference between breastfed infants presenting stools of a more liquid consistency (**Table S2**), although these differences ceased to be significant at visit 5 (see **table S2**). Likewise, the daily number of stools was higher in breastfed infants at visits 1, 2 and 3, but these differences ceased to be significant at visits 4 and 5 (**Table 7**).

**Table 7.** The daily number of stools.

| Number of daily depositions | Visit 1 | Visit 2 | Visit 3 | Visit 4 | Visit 5 |
| --- | --- | --- | --- | --- | --- |
| BFD |  |  |  |  |  |
| Mean (SD) | 5.2 (3.2) | 3.1 (2.3) | 2.3 (1.5) | 2.0 (1.4) | 1.8 (1.0) |
| Median [Min, Max] | 5 [3, 8] | 3 [1, 4] | 2 [1, 3] | 2 [1, 2] | 2 [1, 2] |
| STD |  |  |  |  |  |
| Mean (SD) | 2.5 (2.0) | 1.5 (0.7) | 1.3 (0.6) | 1.5 (1.0) | 1.9 (0.9) |
| Median [Min, Max] | 2 [1, 4] | 1 [1, 2] | 1 [1, 2] | 1 [1, 2] | 2 [1, 2] |
| INN |  |  |  |  |  |
| Mean (SD) | 2.3 (1.4) | 1.6 (0.8) | 1.6 (0.9) | 1.6 (0.9) | 1.9 (0.8) |
| Median [Min, Max] | 2 [1, 3] | 1 [1, 2] | 1 [1, 2] | 1 [1, 2] | 2 [1, 2] |

Data are expressed as mean and maximum and minimum

**Figure 1** shows the tabulation of the different categories of digestive tolerance between every two visits. An increase in the percentage of children with high tolerance is seen throughout the study without significant differences between the different formulations and compared with the BFD group.

**Figure 1.**
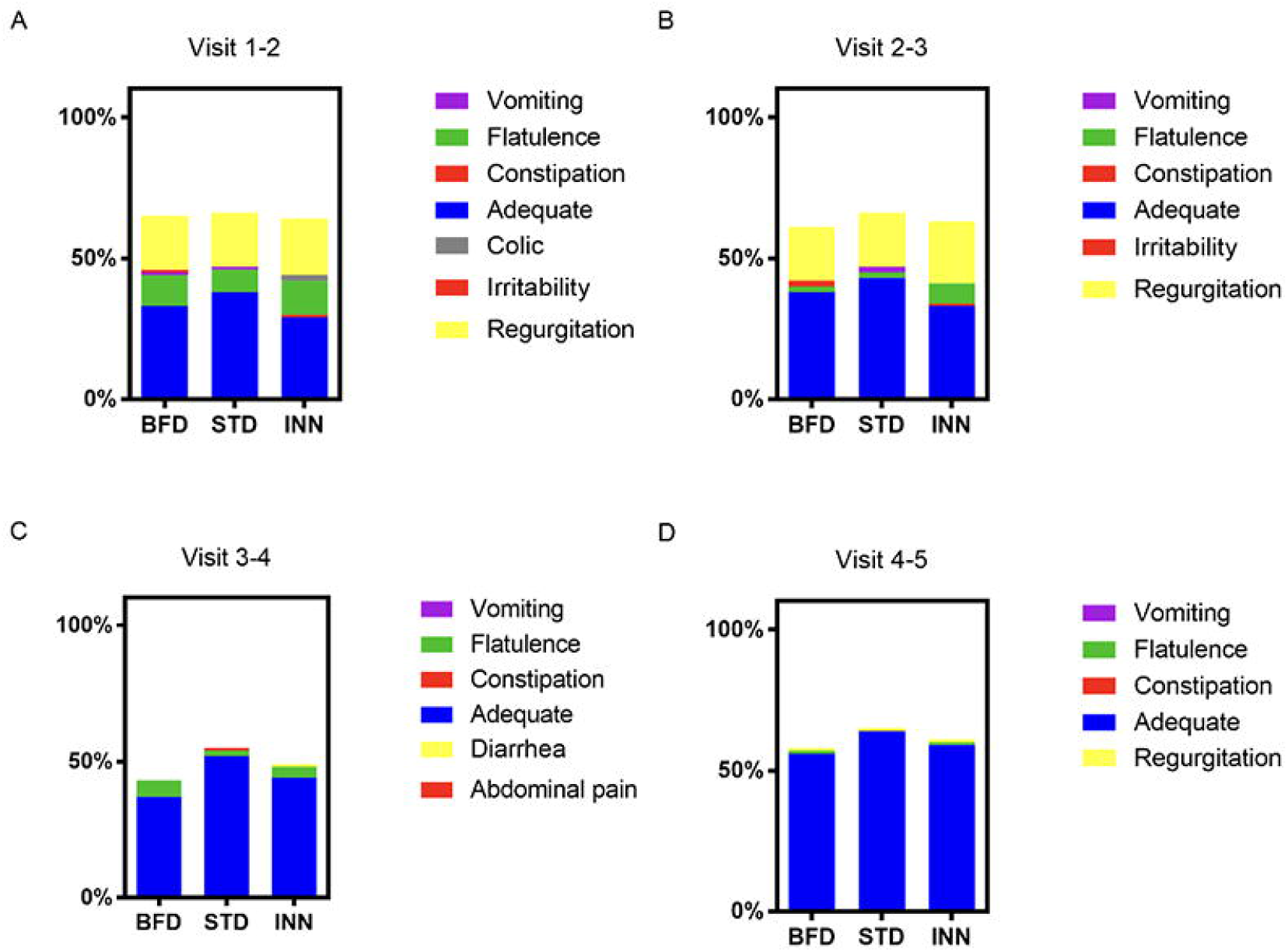
Digestive tolerance. Data are presented as percentages (%). BFD, breastfeeding group; STD, standard group; INN, INNOVA group.

The infant’s behavior was re-coded as altered mood or pleasant mood according to the guardians or parents. **Table S3** shows how the percentage of infants with altered behavior decreased throughout the study. No significant differences were observed among the three ways of feeding.

The study found no differences in tolerability between the different groups (**Table S4**). According to the safety of the different formulations, the majority of the adverse events (n=754) were mild – Grade 1 (94.3%), 13 were moderate-Grade 2 (1.6% of the total) and none were severe-Grade 3. The intensity was not recorded for 33 of the events (4.1%) (**Table 8**). There were no differences between the different groups concerning the intensity of the events. For example, there were 93.0% of mild events in the BFD group, 91.8% in the INN group, and 97.6% in the STD group.

**Table 8.** Gastrointestinal symptoms across the study

| Total adverse events | BFD (N=70) | STD (N=70) | INN (N=70) |
| --- | --- | --- | --- |
| Subjects with at least one adverse event (%) | 62 (88.6%) | 57 (81.4%) | 61 (87.1%) |
| Total number | 227 | 291 | 282 |
| Mild - Grade 1 | 211 | 284 | 259 |
| Moderate - Grade 2 | 3 | 3 | 7 |
| Severe - Grade 3 | 0 | 0 | 0 |
| Potentially fatal - Grade 4 | 0 | 0 | 0 |
| Lethal - Grade 5 | 0 | 0 | 0 |
| Unknown | 13 | 4 | 16 |
| **Adverse events related to feeding** | **BFD (N=70)** | **STD (N=70)** | **INN (N=70)** |
| Subjects with at least one adverse event (%) | 1 (1.4%) | 1 (1.4%) | 1 (1.4%) |
| Total number | 1 | 2 | 2 |
| Mild - Grade 1 | 1 | 2 | 2 |
| Moderate -Grade 2 | 0 | 0 | 0 |
| Severe - Grade 3 | 0 | 0 | 0 |
| Potentially fatal - Grade 4 | 0 | 0 | 0 |
| Lethal - Grade 5 | 0 | 0 | 0 |
| Unknown | 0 | 0 | 0 |

Data are expressed as counts and percentages

Most of the gastrointestinal symptoms were mild, with 1 growth failure, within the BFD group that led to the introduction of artificial breastfeeding together with breastfeeding, two events, neonatal constipation, and abdominal pain, recorded by one infant within the INN group, led to the change of formula and the abandonment of the study and two (infantile colic and infant reflux) recorded by one infant within the STD group that led to the change of formula and the abandonment of the study (**Table 8**).

For morbidities that occurred with a frequency greater than 1%, BFD infants exhibited the lowest incidence and infants fed INN formula experienced significantly fewer general disorders and disturbances compared with the STD group. Indeed, the infants who were fed STD formula had a significantly higher incidence of atopic dermatitis, bronchitis and bronchiolitis events than the infants who were fed BFD or INN formula. (**Table 9**).

**Table 9.** Morbidities that occur with a frequency greater than 1%.

| MedDRA dictionary version 21.0<br>(MedDRA, 2018) | BFD (N=70) |  | STD (N=70) |  | INN (N=70) |  |
| --- | --- | --- | --- | --- | --- | --- |
|  | Case<br>s | % | Cases | % | Cases | % |
| Cold | 42 | 18.50 | 47 | 16.15 | 38 | 13.48 |
| Pyrexia | 16 | 7.05 | 24 | 8.25 | 11 | 3.90 |
| Bronchitis/Bronchiolitis* | 13 | 5.7 | 34 | 11.7 | 17 | 6.00 |
| Upper respiratory tract infection | 11 | 4.85 | 11 | 3.78 | 15 | 5.32 |
| Infantile colic | 14 | 6.17 | 9 | 3.09 | 10 | 3.55 |
| Conjunctivitis | 5 | 2.20 | 11 | 3.78 | 15 | 5.32 |
| Neonatal constipation | 6 | 2.64 | 10 | 3.44 | 9 | 3.19 |
| Acute otitis media | 4 | 1.76 | 6 | 2.06 | 13 | 4.61 |
| Rhinorrhea | 9 | 3.96 | 5 | 1.72 | 6 | 2.13 |
| Gastroenteritis | 4 | 1.76 | 8 | 2.75 | 8 | 2.84 |
| Cough | 9 | 3.96 | 7 | 2.41 | 5 | 1.77 |
| Vaccination complication | 1 | 0.44 | 10 | 3.44 | 7 | 2.48 |
| Laryngitis | 1 | 0.44 | 9 | 3.09 | 7 | 2.48 |
| Atopic dermatitis* | 1 | 0.44 | 11 | 3.78 | 2 | 0.71 |
| Seborrheic dermatitis | 2 | 0.88 | 4 | 1.37 | 5 | 1.77 |
| Neonatal diarrhea | 2 | 0.88 | 3 | 1.03 | 5 | 1.77 |
| Oral candidiasis | 4 | 1.76 | 3 | 1.03 | 2 | 0.71 |
| Nasopharyngitis | 1 | 0.44 | 2 | 0.69 | 6 | 2.13 |
| Infantile eczema | 1 | 0.44 | 5 | 1.72 | 2 | 0.71 |
| Diaper dermatitis | 2 | 0.88 | 2 | 0.69 | 4 | 1.42 |
| Rash | 3 | 1.32 | 2 | 0.69 | 2 | 0.71 |
| Neonatal disorder | 2 | 0.88 | 1 | 0.34 | 4 | 1.42 |
| Upper respiratory tract infection | 2 | 0.88 | 1 | 0.34 | 3 | 1.06 |
| Wheezing | 2 | 0.88 | 3 | 1.03 | 1 | 0.35 |
| Infant reflux | 1 | 0.44 | 2 | 0.69 | 3 | 1.06 |
| Otitis media | 4 | 1.76 | 1 | 0.34 | 1 | 0.35 |
| Gastroesophageal reflux disease | 4 | 1.76 | 0 | 0.00 | 1 | 0.35 |
| Heart murmur | 4 | 1.76 | 1 | 0.34 | 0 | 0.00 |
| Diarrhea | 1 | 0.44 | 0 | 0.00 | 4 | 1.42 |
| Atopic Dermatitis* | 1 | 0.44 | 3 | 1.03 | 0 | 0.00 |
| Traumatic head injury | 0 | 0.00 | 1 | 0.34 | 4 | 1.42 |
| Abdominal pain | 0 | 0.00 | 1 | 0.34 | 3 | 1.06 |
| Bronchial hyperreactivity | 0 | 0.00 | 3 | 1.03 | 0 | 0.00 |
| Decreased appetite | 0 | 0.00 | 3 | 1.03 | 0 | 0.00 |
| Pharyngotonsillitis | 0 | 0.00 | 1 | 0.34 | 3 | 1.06 |
| Umbilical hernia | 0 | 0.00 | 1 | 0.34 | 3 | 1.06 |
| Viral rash | 0 | 0.00 | 1 | 0.34 | 3 | 1.06 |
| Traumatic injury to the mouth | 1 | 0.44 | 0 | 0.00 | 3 | 1.06 |
| Total |  | 173 |  | 246 |  | 225 |

Data are expressed as counts and percentages, \*p<0.05 BFD and INN vs. STD group. For the analysis, the Chi-square test was used with the Yates correction where appropriate. Bronchitis/bronchiolitis showed a p-value of 0.029 (BFD vs STD), 1.0 (BFD vs INN), and 0.026 (STD vs INN). Atopic dermatitis exhibited a p-value of 0.027 (BFD vs STD), 1.0 (BFD vs INN), and 0.029 (STD vs INN)

## 4. Discussion

The present randomized, double-blind, placebo-controlled clinical trial was designed to determine whether a novel starting infant formula with reduced protein content, lower casein to whey protein ratio by increasing the content of α-lactalbumin, influenced weight gain and body composition compared to a standard formula at 6 and 12 months of age. Furthermore, this product contains higher levels of DHA and ARA, as well as a postbiotic thermally inactivated (BPL1™ HT), compared with standard infant formula. An exclusively breastfed population was followed-up as a reference. At 6 and 12 months, infants receiving either INN or STD formula gained more weight than the BFD group, while no difference was observed between STD and INN formulas. BMI was also higher in infants fed both formulas than in those breastfed. Regarding body composition, length, head circumference, and tricipital/subscapular skinfolds, we found that all of the measures were similar between all groups. It is important to note that the INN formula was considered safe based on weight gain and body composition, which were within the normal limits, according to WHO standards. Compared to both formulas, BFD produced stools that were more liquid in consistency. Throughout the study period, all groups exhibited similar digestive tolerance and infant behavior. In terms of the total number of adverse events reported, the STD formula had the highest number, followed by the INN formula and finally, the BFD group. However, the majority of them were not related to the type of feeding. Besides, infants fed either BFD or the INN formula exhibited significantly lower episodes of atopic dermatitis, bronchitis and bronchiolitis events than those fed the STD formula.

In addition to exposure to metabolic and endocrine factors during pregnancy (Monasta et al., 2011), protein intake in formula-fed infants during the first year of life may have a significant impact on growth later in life and the risk of obesity and metabolic disorders in adulthood (Koletzko et al., 2009;Zheng et al., 2018). Indeed, the protein content of infant formula is usually greater than that of human milk to provide all essential amino acids in adequate quantities (Davis et al., 2008). Scientific evidence indicates that infants fed a formula containing more protein gain more weight during the first year of life and are heavier at 2 years of age than infants consuming a formula containing less protein (Koletzko et al., 2009), which reduces the risk of obesity at school age (Weber et al., 2014). In our RCT, infants fed either INN or STD formula gained more weight than the breastfed group at 6 months as measured by differential daily weight gain per day (g). Similar results have been reported in other earlier studies (Baird et al., 2008) even with a relatively low content of protein (Timby et al., 2014). However, previous studies on formulas with a reduced protein-energy ratio of up to 1.8 g/100 kcal have shown more modest differences in growth patterns compared with infants breastfed, similar ot our findings (Räihä et al., 2002;Koletzko et al., 2009;Ren et al., 2022), which supports the hypothesis that the protein-energy ratio plays a key role for weight gain. In our study the intake of INN formula was slightly higher than the STD formula, thus compensating for the lower energy content and protein of the former. Compensation for lower energy density and protein has been reported in other studies (Timby et al., 2014). After 12 months, the STD group was significantly higher than BFD and INN groups. We should note that the INN formula contains 8% less protein per 100 kcal than the STD. Thus, in the first six to eight weeks of life, there is almost no difference between breast milk and formula-fed infants in terms of growth (gain in weight and length). Indeed, it has been previously reported that formula-fed infants gained weight and length more rapidly than breast-fed infants from about two months of age to the end of their first year of life (Ziegler, 2006). Interestingly, the results of a recent review suggest that the difference in weight gain between formula-fed and breastfed infants is relatively small, comparable, and significantly less than the nutritionally significant differences used in determining sample size (Wallingford and Barber, 2019).

In addition to the total protein content, the whey/casein ratio in infant formulas seems to be important for the first year of life. Whey proteins, particularly β-lactoglobulin and α-lactalbumin, are rapidly digested and participate in the building of muscle mass (Boirie et al., 1997), and particularly, α - lactalbumin is the major protein in human milk (Liao et al., 2017). Thus, the addition of bovine α-lactalbumin to infant formula modifies the plasma amino acid pattern of the receiver infant, allowing a reduction in the protein content of the formula. On the other hand, casein proteins are water-insoluble high molecular weight molecules that are a source of phosphate and calcium in human milk, because of the highly phosphorylated nature of β-casein and α-S1-casein, and the requirement for calcium in forming the aggregates of casein micelles (Poth et al., 2008). Nevertheless, the amount should be controlled to avoid improper digestion in early life. In this context, using cow’s milk as the protein source in infant formula might result in an α-casein-dominant formula, which differs from the whey protein dominance in human milk. Further, bovine whey contains a high concentration of β-lactoglobulin, which is absent in human milk (Jackson et al., 2004), therefore, adding bovine α-lactalbumin to infant formula makes it more similar to that of breastfed infants (Lönnerdal and Lien, 2003). Moreover, α-lactalbumin is relatively rich in tryptophan, which may result in satisfactory growth and plasma tryptophan levels similar to those of breastfed infants and infants fed standard formula (Sandström et al., 2008). Nevertheless, in our study, both formula-fed infants gained weight more rapidly compared with exclusively breastfed infants, and there was no significant difference between the groups in terms of length, head circumference, tricipital and subscapular skinfolds, and upper arm circumference, suggesting that both formulas causes similar body composition in infants.

The proportion of fatty acids present in infant formulas is important to ensure the correct growth and development of infants. In the last 20 years, formulas have been supplemented with LC-PUFA in amounts similar to breast milk. Despite the new EU regulation that indicates that AA is not needed to be included, intervention studies assessing the impact of DHA- and AA-supplemented formulas have resulted in numerous positive developmental outcomes (closer to breast-fed infants) including measures of specific cognition functions, visual acuity, and immune responses (Forsyth et al., 2017;Jasani et al., 2017;Lien et al., 2018). AA has different biological functions compared to DHA, for example, AA has exclusive functions in the vasculature and specific aspects of immunity. Undeniably, most of the trials include both DHA and AA, and test development specific to DHA such as neural and visual development. DHA suppresses membrane AA concentrations and its function. Infant formula with DHA and no AA runs the risk of cardio and cerebrovascular morbidity and even mortality through suppression of the favorable oxylipin derivatives of AA (Crawford et al., 2015). International expert consensus suggests that infant formulas that contain DHA at a concentration of 0.3% to 0.5% by weight of total fat and AA at a minimum level equivalent to the amount of DHA should be clinically tested (Koletzko et al., 2020;Campoy et al., 2021). In this study, we used an advanced formula enhanced with a double amount of DHA/AA in comparison with a STD formula to provide evidence about weight gain and body composition, among other outcomes. Throughout the study, the percentage of children with high tolerance increased without significant differences between formulations and compared to the group receiving BFD. According to the guardians or parents, the infant’s behavior was classified as altered mood or pleasant mood. There was a decrease in the number of infants with altered behavior throughout the study. The three methods of feeding did not show any significant differences.

The INN formula was also supplemented with a thermally inactivated postbiotic *Bifidobacterium animalis* subsp. *lactis*, BPL1™ (Carreras et al., 2018;Nataraj et al., 2020), which may confer some benefits regarding body composition, metabolism, and gut microbiota composition. Probiotics have many health benefits by modulating the gut microbiome; nonetheless, techno-functional limitations have made it gradually shift from viable probiotic bacteria towards non-viable postbiotics, paraprobiotics and/or probiotics-derived biomolecules, so-called postbiotics (Nataraj et al., 2020). Since the experimental formula of the present study was supplemented with BPL1™ HT, it should be noted that *in vivo* studies reported that this strain can be considered a Generally Recognized as Safe (GRAS) substance. In terms of the intensity of GI symptoms, tolerability and safety, there were no differences between the different groups (BFD, STD and INN formula). In different contexts and using different methods, infants are fed a variety of products. The formula for infants is available in many variations, many of which are superior to other methods of feeding and some of which are inferior to others. On the other hand, there was a significant difference between BFD group presenting stools of a more liquid consistency, although this difference no longer existed at visit five (12 months). Additionally, BFD infants had more stools per day at visits 1, 2, and 3, but these differences ceased to be significant at visits 4 and 5. STD formula group reported the most of GI symptoms, followed by the INN formula and BFD group. There were no differences in safety across the study between the different groups. Several differences were observed between the two formulas, showing a significant reduction in general disorders and disturbances among children who received the INN formula. STD formula-fed infants were more likely to cause atopic dermatitis, bronchitis, and bronchiolitis than BFD or INN formula-fed infants. Eczema or atopic dermatitis is a common chronic inflammatory skin disease, mostly occurring in children. Indeed, a meta-analysis showed that probiotic supplementation during both the prenatal and the postnatal period reduced the incidence of atopic dermatitis in infants and children, suggesting that starting probiotic treatment during gestation and continuing through the first 6 months of the infant’s life may be of benefit in the prevention of atopic dermatitis (Li et al., 2019). In this context, a study demonstrated that prenatal and postnatal supplementation with a mixture of *B. bifidum* BGN4, *B. lactis* AD011, and *L. acidophilus* AD031 is an effective approach in preventing the development of eczema in infants at high risk of allergy during the first year of life (Kim et al., 2010). Hence, the lower incidence of atopic dermatitis observed in the INN formula group compared to the STD group might be mediated by the supplementation of the postbiotic BPL1™ HT. Likewise, respiratory tract infections represent one of the main health problems in children of different ages (Vesa et al., 2001). Finally, changes in the intestinal microbiota will need to be assessed.

There are some limitations for this study. There were no significant correlations between the type of feeding and the number of adverse events. Nevertheless, the number of infants in the present study was calculated for the growth as the main variable and not for the morbidity.

In conclusion, this clinical trial involved the evaluation of a novel infant formula with reduced content of total protein and modification of the whey/casein ratio by increasing the content of α-lactalbumin, increased levels of both AA and DHA, and postbiotic in comparison with standard infant formula. For exploratory analysis, a third unblinded group of breastfed infants was used. Both formulas gained more weight at 6 and 12 months than the BFD group, while no differences were observed between STD and INN formulas. Infants fed both formulas had a higher BMI than those fed BFD. Body composition, head circumference, and tricipital/subscapular skinfolds were similar between the two groups. We should note that the INN formula is safe and it showed a reduction in atopic dermatitis, bronchitis, and bronchiolitis in infants compared to STD formula. The consistency of the stools produced by BFD was more liquid in comparison with both formulas. For further analysis, it would be necessary to examine more precise health biomarkers and to carry out long-term longitudinal studies.

## Data Availability

All data produced in the present study are available upon reasonable request to the authors

## Author Contributions

AGG, IG and MM were responsible for a careful review of the protocol, design, and methodology. FJRO, JPD and AG provided continuous scientific advice for the study and the interpretation of the results. These authors also wrote and critically reviewed the manuscript. All authors approved the final version of the manuscript.

## Acknowledgments

We thank to the pediatrician team who participated in this study: Alba Corrales; Alfonso Carmona; Amalia López; Ana Isabel Rodriguez; Ana Maderuelo; Ana Terrén-Lora; Eduardo Ortega; Esther Martin; Eva Castillo; Isabel Mayordomo; José Fernando Ferreira; José María Aguilar Diosdado; Maite Santos-Garcia; Mari Carmen Pino Zambrano; María José Carnicero; Maria Teresa Santos; Vasilica Doina and Xavier Riopedre. Julio Plaza-Diaz is part of the “UGR Plan Propio de Investigación 2016” and the “Excellence actions: Unit of Excellence on Exercise and Health (UCEES), University of Granada”. Julio Plaza-Diaz is supported by a fellowship awarded to postdoctoral researchers at foreign universities and research centers from the “Fundación Ramón Areces”, Madrid, Spain.

## Supplementary Material

Supplementary material is expressed as tables and is found in the supplemtary file (Tables S1-S4).

## Data Availability Statement

The datasets used and/or analyzed during the current study are available from the corresponding author upon reasonable request.

