## Supplemental material for "Effects of a novel infant formula on weight gain and body composition of infants: The INNOVA 2020 study"

**Table S1.** Body mass index, weight, and height percentiles.

| Body mass index percentiles | | | | **P-value** | | |
| --- | --- | --- | --- | --- | --- | --- |
| **Visits** | **BFD** | **STD** | **INN** | **Formula** | **Visit** | **Formula x visit** |
|  | **(N=58)** | **(N=65)** | **(N=62)** |  |  |  |
| **Visit 1** | 0.45 ± 0.26 | 0.42 ± 0.24 | 0.47 ± 0.25 | 0.504 | 0.009 | 0.006 |
| **Visit 2** | 0.45 ± 0.25 | 0.43 ± 0.25 | 0.41 ± 0.26 |  |  |  |
| **Visit 3** | 0.40 ± 0.24 | 0.44 ± 0.29 | 0.50 ± 0.29 |  |  |  |
| **Visit 4** | 0.43 ± 0.26 | 0.47 ± 0.31 | 0.55 ± 0.31 |  |  |  |
| **Visit 5** | 0.51 ± 0.28 | 0.57 ± 0.31 | 0.55 ± 0.31 |  |  |  |
| Height percentiles | | | | **P-value** | | |
| **Visits** | **BFD** | **STD** | **INN** | **Formula** | **Visit** | **Formula x visit** |
|  | **(N=58)** | **(N=65)** | **(N=62)** |  |  |  |
| **Visit 1** | 0.54 ± 0.30 | 0.43 ± 0.30 | 0.48 ± 0.26 | 0.433 | 0.011 | <0.001 |
| **Visit 2** | 0.58 ± 0.33 | 0.52 ± 0.30 | 0.60 ± 0.27 |  |  |  |
| **Visit 3** | 0.52 ± 0.31 | 0.55 ± 0.30 | 0.59 ± 0.27 |  |  |  |
| **Visit 4** | 0.47 ± 0.30 | 0.52 ± 0.31 | 0.56 ± 0.26 |  |  |  |
| **Visit 5** | 0.53 ± 0.31 | 0.58 ± 0.30 | 0.63 ± 0.27 |  |  |  |
| Weight percentiles | | | | **P-value** | | |
| **Visits** | **BFD** | **STD** | **INN** | **Formula** | **Visit** | **Formula x visit** |
|  | **(N=58)** | **(N=65)** | **(N=62)** |  |  |  |
| **Visit 1** | 0.49 ± 0.27 | 0.40 ± 0.27 | 0.47 ± 0.25 | 0.368 | <0.001 | <0.001 |
| **Visit 2** | 0.51 ± 0.26 | 0.45 ± 0.27 | 0.49 ± 0.25 |  |  |  |
| **Visit 3** | 0.43 ± 0.26 | 0.47 ± 0.30 | 0.54 ± 0.26 |  |  |  |
| **Visit 4** | 0.42 ± 0.25 | 0.48 ± 0.32 | 0.56 ± 0.28 |  |  |  |
| **Visit 5** | 0.53 ± 0.26 | 0.59 ± 0.32 | 0.61 ± 0.29 |  |  |  |

Data are expressed as mean and standard deviation

**Table S2.** Stool Characteristics.

|  | Visit 1 | | | Visit 2 | | | Visit 3 | | | Visit 4 | | | Visit 5 | | |
| --- | --- | --- | --- | --- | --- | --- | --- | --- | --- | --- | --- | --- | --- | --- | --- |
| Stool consistency | **BFD** | **STD** | **INN** | **BFD** | **STD** | **INN** | **BFD** | **STD** | **INN** | **BFD** | **STD** | **INN** | **BFD** | **STD** | **INN** |
| Soft | 3 (4.3%) | 22 (31.4%) | 12 (17.1%) | 5 (7.7%) | 17 (25.8%) | 12 (18.8%) | 8 (13.1%) | 16 (24.2%) | 13 (20.6%) | 3 (5.2%) | 16 (24.2%) | 12 (19.0%) | 11 (19.0%) | 13 (20.0%) | 10 (16.1%) |
| Hard | 5 (7.1%) | 6 (8.6%) | 11 (15.7%) | 0 (0.0%) | 1 (1.5%) | 3 (4.7%) | 0 (0.0%) | 0 (0.0%) | 1 (1.6%) | 1 (1.7%) | 4 (6.1%) | 0 (0.0%) | 4 (6.9%) | 7 (10.8%) | 5 (8.1%) |
| Liquid | 27 (38.6%) | 4 (5.7%) | 7 (10.0%) | 24 (36.9%) | 0 (0.0%) | 0 (0.0%) | 13 (21.3%) | 0 (0.0%) | 1 (1.6%) | 9 (15.5%) | 1 (1.5%) | 4 (6.3%) | 0 (0.0%) | 0 (0.0%) | 0 (0.0%) |
| Normal | 0 (0.0%) | 3 (4.3%) | 2 (2.9%) | 0 (0.0%) | 4 (6.1%) | 1 (1.6%) | 2 (3.3%) | 7 (10.6%) | 3 (4.8%) | 2 (3.4%) | 8 (12.1%) | 2 (3.2%) | 7 (12.1%) | 9 (13.8%) | 16 (25.8%) |
| Pasty | 8 (11.4%) | 31 (44.3%) | 34 (48.6%) | 13 (20.0%) | 40 (60.6%) | 45 (70.3%) | 13 (21.3%) | 38 (57.6%) | 38 (60.3%) | 23 (39.7%) | 33 (50.0%) | 42 (66.7%) | 36 (62.1%) | 36 (55.4%) | 31 (50.0%) |
| Semiliquid | 27 (38.6%) | 4 (5.7%) | 4 (5.7%) | 23 (35.4%) | 4 (6.1%) | 3 (4.7%) | 25 (41.0%) | 5 (7.6%) | 7 (11.1%) | 20 (34.5%) | 4 (6.1%) | 3 (4.8%) | 0 (0.0%) | 0 (0.0%) | 0 (0.0%) |

Data are expressed as counts and percentages

### Table S3. Infant’s behavior

| Visit |  | BFD | STD | INN |
| --- | --- | --- | --- | --- |
| 1 | N | 70 | 70 | 70 |
|  | Altered mood | 27 (38.6%) | 29 (41.4%) | 22 (31.4%) |
|  | Good mood | 43 (61.4%) | 41 (58.6%) | 48 (68.6%) |
| 2 | N | 65 | 66 | 64 |
|  | Altered mood | 22 (33.8%) | 18 (27.3%) | 18 (28.1%) |
|  | Good mood | 43 (66.2%) | 48 (72.7%) | 46 (71.9%) |
| 3 | N | 61 | 66 | 63 |
|  | Altered mood | 16 (26.2%) | 9 (13.6%) | 13 (20.6%) |
|  | Good mood | 45 (73.8%) | 57 (86.4%) | 50 (79.4%) |
| 4 | N | 58 | 66 | 63 |
|  | Altered mood | 13 (22.4%) | 13 (19.7%) | 7 (11.1%) |
|  | Good mood | 45 (77.6%) | 53 (80.3%) | 56 (88.9%) |
| 5 | N | 58 | 65 | 62 |
|  | Altered mood | 10 (17.2%) | 4 (6.2%) | 6 (9.7%) |
|  | Good mood | 48 (82.8%) | 61 (93.8%) | 56 (90.3%) |
| Total | N | 312 | 333 | 322 |
|  | Altered mood | 88 (28.2%) | 73 (21.9%) | 66 (20.5%) |
|  | Good mood | 224 (71.8%) | 260 (78.1%) | 256 (79.5%) |

Data are expressed as counts and percentages

### Table S4. Tolerability and overall assessment by parents or guardians across the study

| Tolerability | BFD (N=65) | STD (N=66) | INN (N=64) | Overall assessment by parents or guardians | BFD (N=65) | STD (N=66) | INN (N=64) |
| --- | --- | --- | --- | --- | --- | --- | --- |
| Visit 2 | | | | Visit 2 | | | |
| Adequate | 5 (7.7%) | 4 (6.1%) | 1 (1.6%) | Acceptable | 1 (1.5%) | 3 (4.6%) | 2 (3.1%) |
| Good | 50 (76.9%) | 48 (72.7%) | 50 (78.1%) | Good | 54 (83.1%) | 40 (61.5%) | 47 (73.4%) |
| Very good | 7 (10.8%) | 13 (19.7%) | 13 (20.3%) | Very good | 9 (13.8%) | 16 (24.6%) | 14 (21.9%) |
| Data not available | 2 (3.1%) | 0 (0.0%) | 0 (0.0%) | Satisfactory | 0 (0.0%) | 5 (7.7%) | 1 (1.6%) |
| Not satisfactory | 1 (1.5%) | 1 (1.5%) | 0 (0.0%) | Not satisfactory | 1 (1.5%) | 1 (1.5%) | 0 (0.0%) |
| Visit 3 | | | | Visit 3 | | | |
| Tolerability | BFD (N=65) | STD (N=66) | INN (N=64) | Overall assessment by parents or guardians | BFD (N=65) | STD (N=66) | INN (N=64) |
| Adequate | 3 (4.9%) | 5 (7.6%) | 2 (3.2%) | Acceptable | 6 (9.8%) | 9 (13.6%) | 2 (3.2%) |
| Good | 38 (62.3%) | 47 (71.2%) | 37 (58.7%) | Good | 37 (60.7%) | 40 (60.6%) | 40 (63.5%) |
| Very good | 14 (23.0%) | 13 (19.7%) | 23 (36.5%) | Very good | 15 (24.6%) | 13 (19.7%) | 19 (30.2%) |
| Data not available | 3 (4.9%) | 0 (0.0%) | 1 (1.6%) | Satisfactory | 0 (0.0%) | 3 (4.5%) | 2 (3.2%) |
| Not satisfactory | 3 (4.9%) | 1 (1.5%) | 0 (0.0%) | Not satisfactory | 3 (4.9%) | 1 (1.5%) | 0 (0.0%) |
| Visit 4 | | | | Visit 4 | | | |
| Tolerability | BFD (N=58) | STD (N=66) | INN (N=63) | Overall assessment by parents or guardians | BFD (N=58) | STD (N=66) | INN (N=63) |
| Adequate | 2 (3.4%) | 8 (12.1%) | 1 (1.6%) | Acceptable | 2 (3.4%) | 9 (13.6%) | 1 (1.6%) |
| Good | 39 (67.2%) | 44 (66.7%) | 38 (60.3%) | Good | 40 (69.0%) | 44 (66.7%) | 41 (65.1%) |
| Very good | 14 (24.1%) | 13 (19.7%) | 23 (36.5%) | Very good | 14 (24.1%) | 9 (13.6%) | 17 (27.0%) |
| Data not available | 3 (5.2%) | 0 (0.0%) | 0 (0.0%) | Satisfactory | 2 (3.4%) | 4 (6.1%) | 3 (4.8%) |
| Not satisfactory | 0 (0.0%) | 1 (1.5%) | 1 (1.6%) | Not satisfactory | 0 (0.0%) | 0 (0.0%) | 1 (1.6%) |
| Visit 5 | | | | Visit 5 | | | |
| Tolerability | BFD (N=58) | STD (N=65) | INN (N=62) | Overall assessment by parents or guardians | BFD (N=58) | STD (N=65) | INN (N=62) |
| Adequate | 2 (3.4%) | 5 (7.7%) | 0 (0.0%) | Acceptable | 2 (3.4%) | 9 (13.6%) | 1 (1.6%) |
| Good | 33 (56.9%) | 34 (52.3%) | 40 (64.5%) | Good | 40 (69.0%) | 44 (66.7%) | 41 (65.1%) |
| Very good | 18 (31.0%) | 24 (36.9%) | 21 (33.9%) | Very good | 14 (24.1%) | 9 (13.6%) | 17 (27.0%) |
| Data not available | 4 (6.9%) | 0 (0.0%) | 0 (0.0%) | Satisfactory | 2 (3.4%) | 4 (6.1%) | 3 (4.8%) |
| Not satisfactory | 1 (1.7%) | 0 (0.0%) | 1 (1.6%) | Not satisfactory | 0 (0.0%) | 0 (0.0%) | 1 (1.6%) |

Data are expressed as counts and percentages
